# Etiological Group Analysis of Cytokine Response To Influenza-Like Illnesses

**DOI:** 10.1101/2020.12.16.20248297

**Authors:** Marina A. Plotnikova, Sergey A. Klotchenko, Alexey A. Lozhkov, Kirill I Lebedev, Alexander S. Taraskin, Irina L. Baranovskaya, Maria A Egorova, Edward S. Ramsay, Andrey V. Vasin

## Abstract

Respiratory infections, collectively, are one of the world’s most common and serious illness groups. As recent observations have shown, the most severe courses of acute respiratory infection, often leading to death, are due to uncontrolled cytokine production (hypercytokinemia). The research presented is devoted to assessment of mRNA expression of specific cytokines (IL-1b, IL-2, IL-4, IL-6, IL-8, IL-10, IL-18, TNF-α, IFN-λ) and MxA in whole blood leukocytes, by means of real-time PCR. This study involved 364 patients with respiratory illness being treated in clinics in St. Petersburg (Russia) in 2018-2019 and 30 healthy subjects. In 70% of patients, bacterial or viral pathogens were identified, with influenza viral infections (types A, B) prevailing. Cytokine analysis was carried out in the acute phase of illness (2-3 days from onset of initial symptoms) and in the stage of recovery (days 9-10). Significant increases in the expression of IL-18, TNF, and IL-10 were observed, relative to controls, only with influenza viral infections. We have shown a difference in IL-6 mRNA expression in patients with bacterial or viral pathogens. No significant difference was found in WBC IL-4 expression levels between patients and healthy subjects. Investigation of the nuances of systemic cytokine production, in response to specific viral and bacterial pathogens, makes it possible to: assess the risks of developing hypercytokinemia during respiratory infection with agents circulating in the human population; and to predict the pathogenicity and virulence of circulating threats.

## Introduction

Influenza and influenza-like infections (ILIs), both viral and bacterial in origin, are the most common group of acute illnesses. Influenza-like infections are cyclic (seasonal), affect a large proportion of the population, and feature huge epidemic or pandemic potential. Various respiratory pathogens elicit similar symptoms, which can range from mild to severe. The primary cellular target of respiratory viruses is airway epithelium. These cells, in response to infection, secrete excessive levels of interferons (IFNs) and proinflammatory cytokines such as: TNF-α; IL-1b; IL-6; IL-8; and chemokines (CCL2, CCL5, CXCL8, CXCL10) [Kreijtz et al., 2011].

Cytokines are produced by the local cellular environment at sites of infection to promote antiviral activity and to recruit innate immune cells. It has been shown that production of IL-6 and IFN-α by respiratory epithelium is an important factor in the formation of both local (sore throat, cough, runny nose) and systemic (fever, myalgia, fatigue) symptoms of ILI [Hayden et al., 1998]. The recruitment of immune cells to the area of infection triggers secondary cytokine production by blood leukocytes. Systemic cytokine production by white blood cells (WBCs) is of key importance in the development of immunopathological conditions with ILI. Hypercytokinemia (cytokine storm) of some blood cytokines is positively correlated with illness severity measures in outpatients; it is also associated with fatal outcome. Cytokine storms are associated with a wide variety of infectious and noninfectious diseases (Tisoncik et al., 2012; Teijaro, 2017). Infectious agents that cause hypercytokinemia include Epstein-Barr virus, cytomegalovirus, and group A streptococcus. Respiratory pathogens and infections are especially associated with hypercytokinemia, such as: H5N1 influenza (Xi-Zhi and Thomas, 2017); MERS; SARS (Channappanavar&Perlman, 2017); and SARS-CoV-2 (Song et al., 2020). Sustained, elevated cytokine levels have been implicated as a sign of poor COVID-19 prognosis [Tang et al., 2020]. High serum levels of pro-inflammatory cytokines (IFN-γ, IL-1, IL-6, IL-12, TGF-β) and chemokines (CCL2, CXCL10, CXCL9, IL-8) have been noted in SARS patients with severe illness compared to individuals with uncomplicated SARS.

Elevated systemic IL-6 and IL-10 levels have been reported in children with severe lower respiratory tract RSV infections [Russell et al., 2017]. Measurement of plasma cytokines has revealed elevated levels of IL-10, IL-6, and IFN-γ in H5N1-infected individuals. Concentrations of IL-6, IL-10, IL-15, IP-10, IL-2R, HGF, ST2, and MIG have been shown to be significantly higher in patients with severe influenza A(H1N1)pdm09 virus infection, compared with mild influenza A(H1N1)pdm09 virus or rhinovirus infection [Bradley-Stewart et al., 2013]. This study is aimed at analyzing cytokine expression in WBCs of patients with viral or bacterial respiratory infection. Characterization of systemic cytokine responses among ILI patients facilitates our understanding of the host immune response. It may also provide prognostic parameters useful in community-acquired pneumonia diagnostics.

## Materials and methods

### Patient information and selection criteria

The study involved 364 patients with respiratory illness being treated at clinics in St. Petersburg (Russia) in 2018-2019. Inclusion of patients in the non-control group was based on the presence of the following signs of acute respiratory illness: fever; intoxication syndrome (weakness, headache, muscle pain); and/or catarrhal syndrome (nasal congestion, rhinorrhea, sore throat, cough, chest pain). On the second or third day after onset of clinical symptoms, samples (nasal and throat swabs, blood samples for WBC isolation) were collected from patients. Following recovery (10-14 days after onset), blood was again taken from patients for analysis.

The control group consisted of donors, aged 25 to 60 years without diagnosed chronic illness, who were healthy at the time of sampling. Serum specimens were provided by the Blood Transfusion Center, Research Institute of Hematology and Transfusion Science (contract No. 128_15092017). All subjects gave their informed consent for inclusion before participation in the study. The study and its protocols were approved by the Smorodintsev Research Institute of Influenza institutional Ethics Committee (protocol No. 108, dated September 03, 2018).

### Diagnosis of pathogens

Laboratory diagnosis of pathogens in selected swabs was performed by RT-PCR using certified AmpliSens Biotechnologies kits [Guzhov et al., 2020].

### Isolation of white blood cells

Blood for WBC isolation was collected in vacuum tubes with sodium heparin. Eight milliliters of blood, diluted with DPBS to a volume of 12 ml, was introduced (avoiding mixing) into a tube containing 9 ml of Lymphosep (BioWest). Tubes were then centrifuged at 400g for 20 min.; resulting WBC layers were taken and washed twice with DPBS containing 2% FBS. Prior to analysis, frozen cells were stored in liquid nitrogen vapor (RPMI storage medium containing 10% DMSO, 50% FBS).

### RNA isolation and RT-qPCR

Total RNA preparations were extracted using the RNeasy mini kit (QIAGEN). Following RNA extraction, samples were reverse transcribed using M-MLV reverse transcriptase (M-MLV RT) (Promega, USA). A mixture of 1-2 μg total RNA and 0.5 μg oligo(dT)_16_ primers (DNA-Synthesis, Russia), adjusted with ultrapure water to a final volume of 15 μl, was incubated at 70°C for 10 min for pre-annealing. Tubes were immediately cooled on ice, followed by addition of final reaction component mix (all Promega): 4 μl 5x MMLV Reaction Buffer; 0.5 μl 5 mM dNTPs; 200 u M-MLV RT; 25 u RNase inhibitor; and ultrapure water to 10 μl. Complementary DNA synthesis was carried out at 42°C for 60 min; products were stored at ^−^20°C until use. qPCR was performed using the 2x BioMaster HS-qPCR reagent (BioLabMix) and previously-developed primers [Plotnikova et al., 2016]. Absolute expression values were calculated by the ΔCt method using GAPDH and β-actin as normalization genes.

### Statistical analysis

Because variables were not normally distributed, a nonparametric Kruskal-Wallis test was used to identify multiple differences between groups. Dunn’s multiple comparisons test was used for pairwise comparison of patient groups with the healthy volunteer group. Comparison of paired groups was performed using the Wilcoxon matched-pairs signed rack test. A Spearman’s test was used for correlation analysis. Statistical significance was considered based on p-value: p<0.05, two-tailed, were considered to be significant. Statistical analyses were performed using GraphPad Prism 6.0 software (GraphPad Software, USA).

## Results

### General patient characteristics

In total, 364 patients, St. Petersburg (Russia) residents aged 18 to 90, were examined during the 2018-2019 epidemic season. All patients had moderate to severe symptoms typical of influenza-like respiratory infection (ILI) such as runny or congested nose, moderate fever (over to 39°C), myalgia, and/or sore throat. Patient swabs were examined for the presence of: influenza A or B viruses (IVA, IVB); human orthopneumovirus (RSV); human Metapneumovirus (HMPV); human parainfluenza virus types 1-4 (HPIVs); human Coronaviruses (HCoV) that cause common cold (not SARS or MERS); human Rhinovirus (HRV); human Adenovirus serotypes B, C, or E (HAdV); and human Bocavirus. Samples were also analyzed for the presence of the bacterial pathogens *Neisseria meningitidis, Haemophilus influenza*, and *Streptococcus pneumoniae*. RT-PCR analysis of nasal and throat swabs identified a pathogen in 62.91% of patients. More than half (36.26%) were IVA or IVB (Fig. 1A).

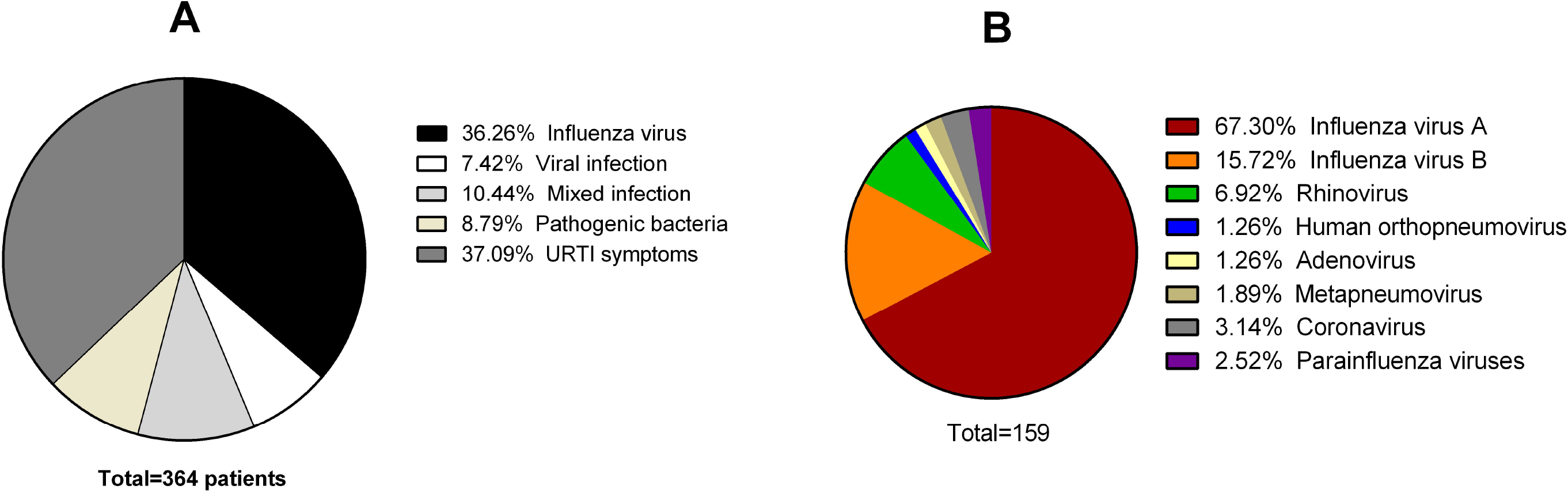

The predominant viral agents, after influenza viruses, were: HRV (6.92%); HCoV (3.14%); and HPIVs (2.52%) (Fig. 1B). In 37% of patients with ILI symptoms, a pathogen could not be detected. Presumably, these patients were ill with whooping cough, *Bordetella bronchiseptica* infection, diphtheria (Corynebacterium), or pneumonia caused by Mycoplasma pneumoniae or Chlamydia pneumoniae. Those specific pathogens were not analyzed. Within 2-3 days from the onset of symptoms, blood was collected from all patients, and WBC-expressed cytokine levels were studied. Some patients were additionally (blood) sampled for post-recovery studies (10-14 days from disease onset). For the control group, 32 healthy volunteers were selected.

### Increased MxA and cytokine mRNA levels in peripheral WBCs

To investigate the role of WBC cytokine production in various ILI etiologies, we analyzed the mRNA expression levels of selected pro-inflammatory (IL-1b, TNF-α, IL-6, IL-1b) and anti-inflammatory (IL-4, IL-10) cytokines. Peripheral WBCs of patients and healthy donors (HD) were used. We found that mRNA levels of MxA, IL-1b, TNF-α, IL-8, and IL-10 were significantly higher in the WBCs from all ILI patient groups compared with healthy controls (Table 1). Expression of IL-18 was significantly different in all patient groups, compared to the control group, with the exception of the heterogeneous group of patients with ILI symptoms of undetermined origin.

**Table 1:**
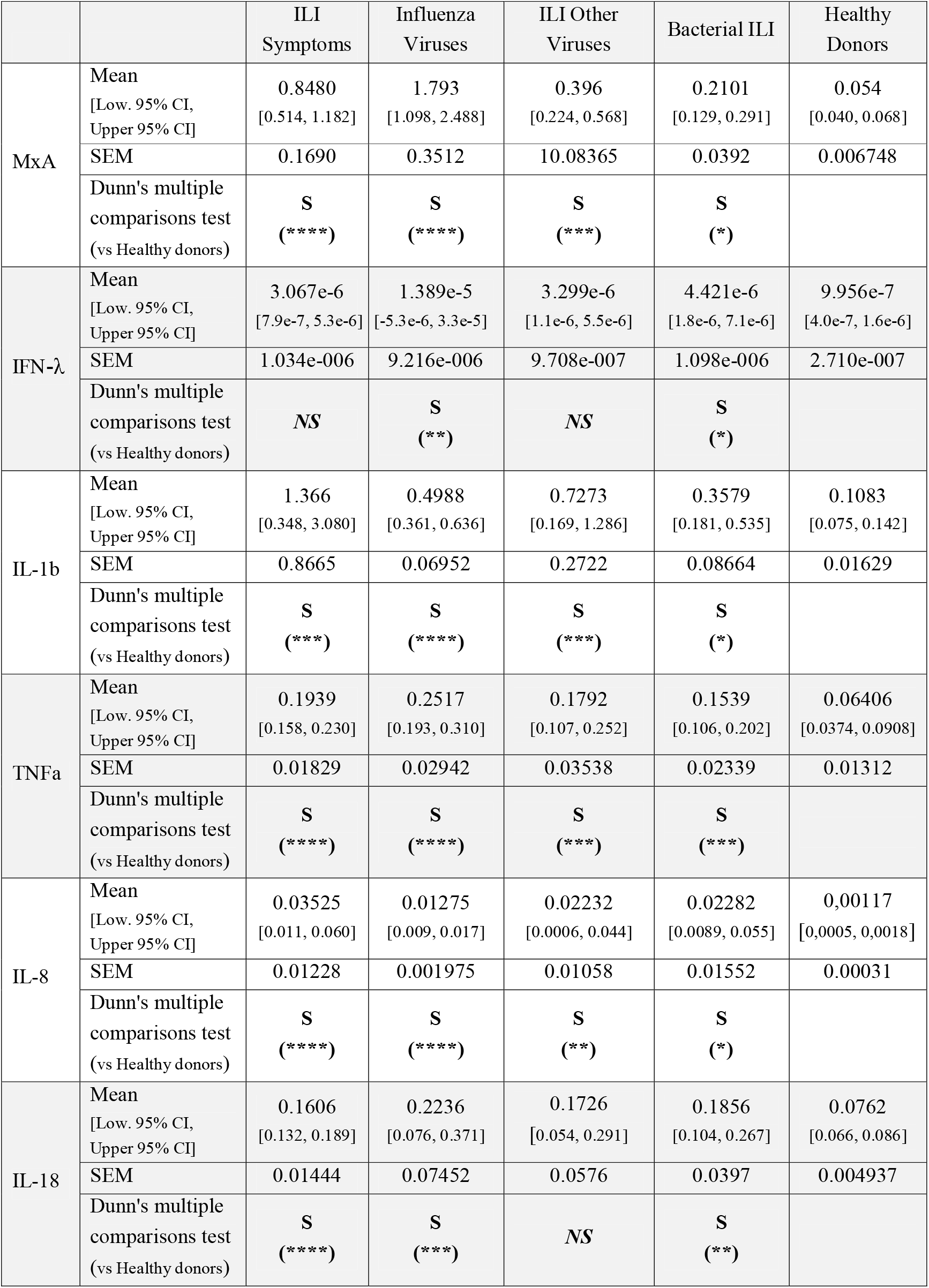
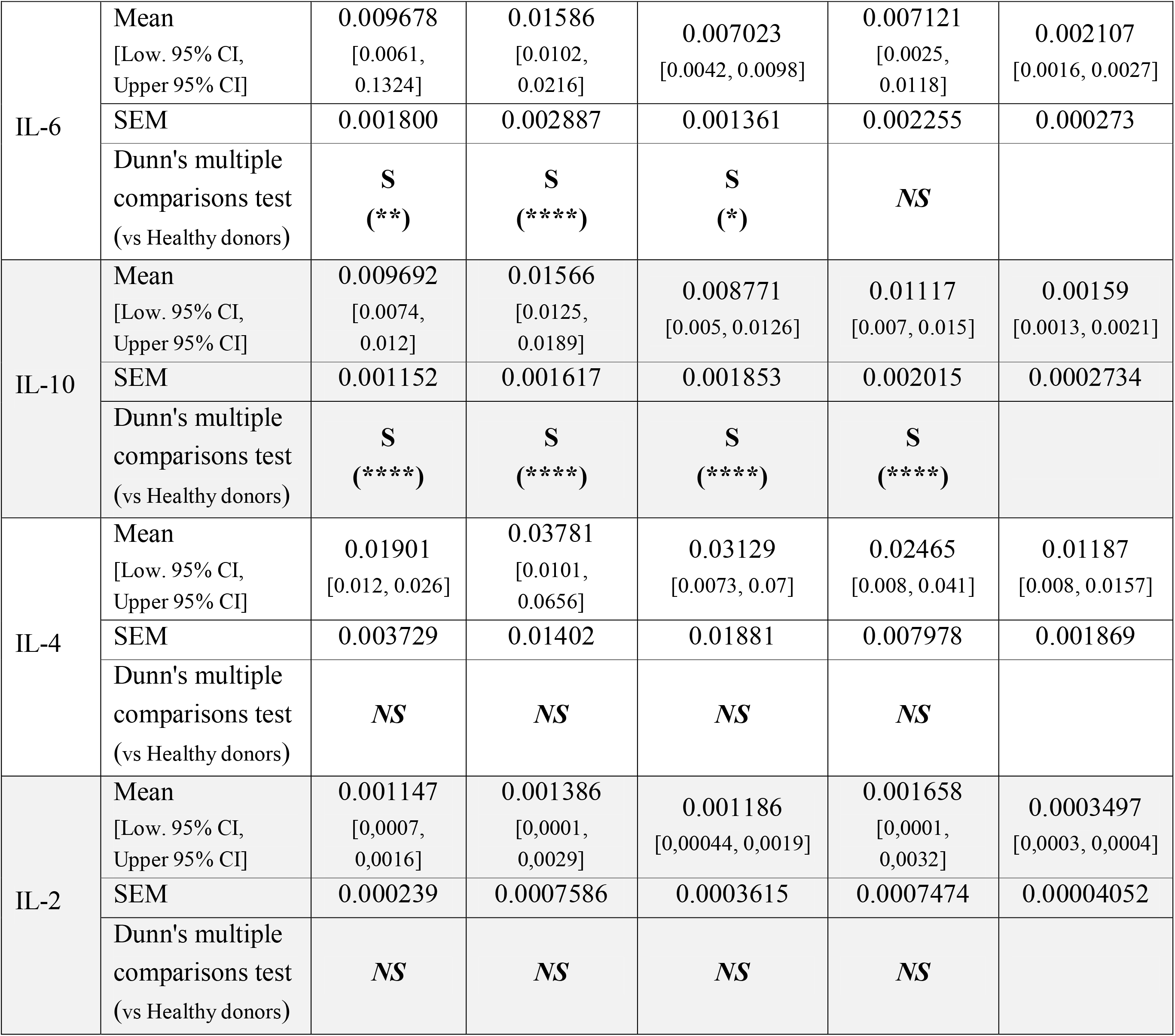
MxA and cytokine differential expression (delta Ct) in patient WBCs.

Interestingly, WBCs also showed increased IFN-λ expression, but only in patients with influenza (approximately 14-fold) or bacterial infection (4-fold). Current literature indicates that: IFN-λ is expressed in DCs, respiratory epithelial cells, keratinocytes, hepatocytes, and others; it largely depends on IRF3 and NF-κB [Hermant&Michiels, 2014]. Differences in IL-2 and IL-4 mRNA expression in all respiratory illness groups were insignificant (p > 0.05) in comparison with the healthy volunteer group.

Interestingly, there were also statistically significant increases in IL-6 expression with viral infection (including influenza), as well as with ILI of unknown origin, compared to the control group. However, analysis suggested no differences between bacterial ILI and healthy donors in terms of WBC IL-6 expression. IL-6 is a key mediator of acute phase inflammation and its overexpression may be the cause of the so-called cytokine storm phenomenon. Peripheral blood IL-6 concentration is used to assess systemic cytokine reaction intensity and to forecast cytokine storm. The absence of significant changes in WBC IL-6 expression in patients with bacterial ILI can be explained by the fact that the IL-6 measured is a secondary response, with production stimulated primarily by TNF and IL-1b (not respiratory pathogens). Increased WBC IL-6 expression may be a sign of transition of the infectious process to the systemic level and the development of a cytokine storm. In the analyzed bacterial infections, cytokine storm is not a typical phenomenon, which is probably why IL-6 level did not change.

All patient groups had increased systemic production of IL-10, which probably provides a negative regulation of systemic response to local respiratory infection. Expression of the interferon-inducible MxA protein, which possesses specific antiviral activities (especially with influenza virus infection), was also increased in patients with bacterial ILI. Presumably, this may be due to the fact that the bacterial infections analyzed were secondary in nature, the trigger for which was a primary respiratory viral infection. However, MxA mRNA levels in patients with influenza virus (both type A and type B) were twice as high as those with bacterial ILI (p < 0.0001). It is possible that MxA expression is further regulated at the translational level with bacterial infection. Since the analyzed patient groups were quite heterogeneous in terms of pathogens, we next compared cytokines in patients with only viral infection.

### Cytokine changes, depending on viral pathogen

The WBC cytokine statuses of patients with ILI caused by different viral pathogens (IVA, IVB, HRV, HCoV, HPIVs) were studied separately. Other viruses caused too few illnesses to form a statistically reliable group. Unfortunately, all the pathogens represented in the groups were RNA-containing viruses. DNA-containing viruses, such as HAdV and human Bocavirus, was found in just two people, with Bocavirus occurring in patients with only a concomitant bacterial infection. ANOVA analysis, using the Kruskal-Wallis test for nonparametric samples, showed no significant differences (WBC expression) in any patient group for: IL-2 vs. control (p = 0.2369); or IL-4 versus control (p = 0.1868). Influenza-related infections, both IVA and IVB, caused significant increases in the levels of MxA, IL-6, IL-8, IL-1b, TNF, IL-10, and IL-18 compared to the control. Interestingly, a significant increase in the expression of IL-18, TNF, and IL-10 was observed only in the case of influenza infection relative to control. Other pathogens caused random changes in these cytokines’ mRNA levels, and differences (comparing these groups of viral infection with the control group or with each other) were unreliable. Further, we showed earlier that cytokine expression in the heterogeneous ILI group was increased approximately 3-fold (TNF) and 6-fold (IL-10), compared to the control group. Pathogens not included in statistical analysis as discrete groups (HAdV, HMPV, RSV, human Bocavirus) appear to have made a large contribution to these changes. IL-18 mRNA levels in the ‘non-influenza viral pathogen’ group did not differ from the control group, which is consistent with the previously obtained results for the heterogeneous ILI group. It is interesting to note that IVB increased IL-6 and IL-8 expression by about 2-fold compared to IVA. In addition, IVB caused statistically significant differences in MxA expression compared to HRV (3.7-fold) and HPIVs (10.5-fold). IVB also increased IL-6 expression, compared to HCoV (12-fold). IFN-λ expression increased only in response to IVB infection. IL-b levels were also significantly increased in patients with HCoV infection, as was IL-8 in patients with HRV infection.

### Cytokine Comparison, Acute Phase Versus Recovery Phase

To assess the formation of a systemic immune response, a cytokine study was also performed 10-14 days after the onset of illness. Since the analyzed ILIs are acute illnesses, pair samples were also taken during the recovery period. Interestingly, on days 10-14 of the study, IL-6 mRNA expression levels in patient WBCs were similar to those measured at the first point. Like early samples (days 2-3), late samples (days 10-14) showed increased IL-6 levels (compared with controls) in all patients, with the exception of the ‘bacterial infection’ group. Interleukin-6 probably manifests its regulatory properties in the recovery phase analyzed: suppressing TNF and IL-1b secretion; eliciting increased IL-2 production by T-helper cells; and facilitating switching from IgM to IgG. This is also evidenced by a significant decrease in the levels of MxA and certain proinflammatory cytokines (IL-1b, TNF) by 10-14 days of infection with influenza and heterogeneous ILIs (Fig. 3).

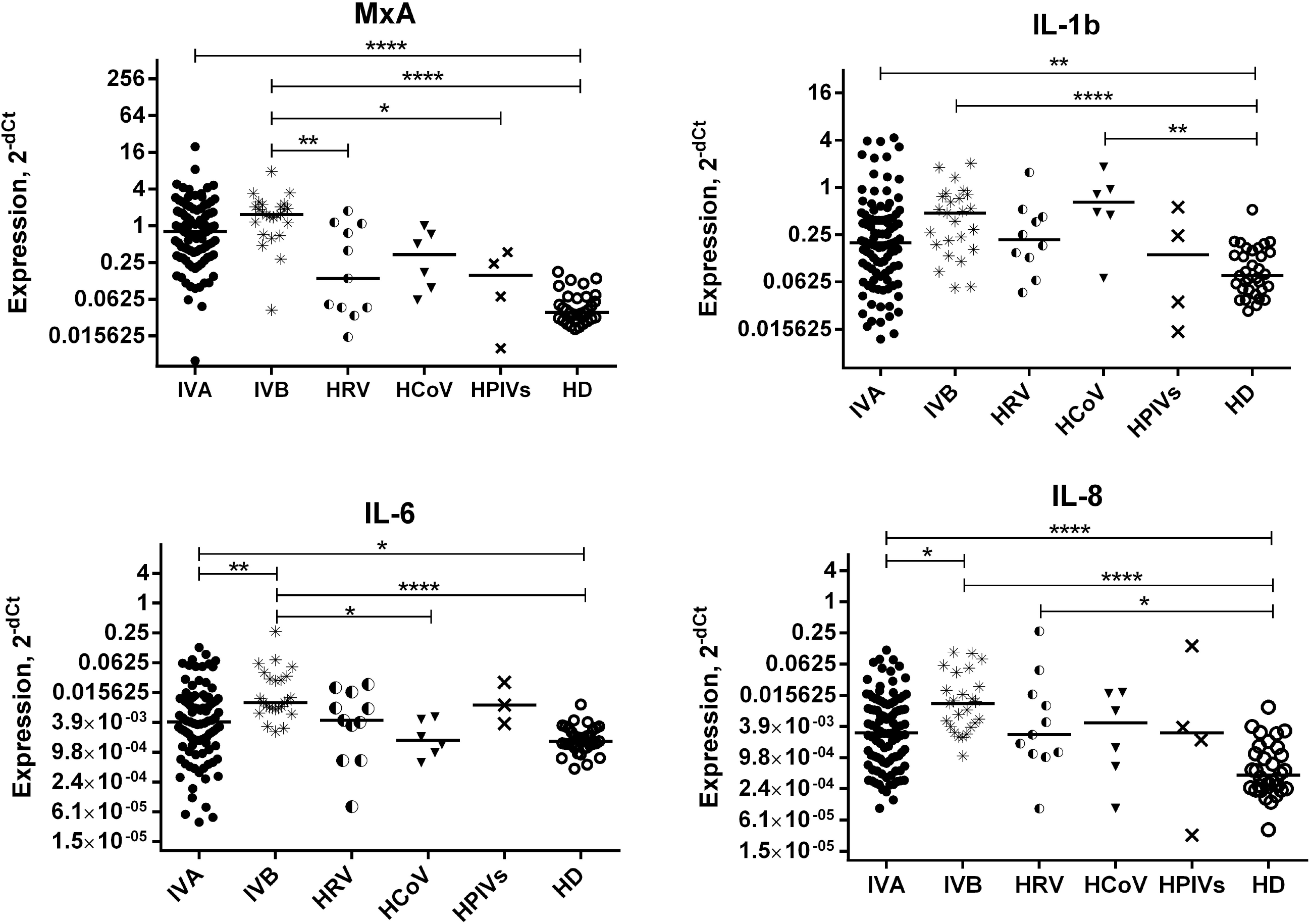

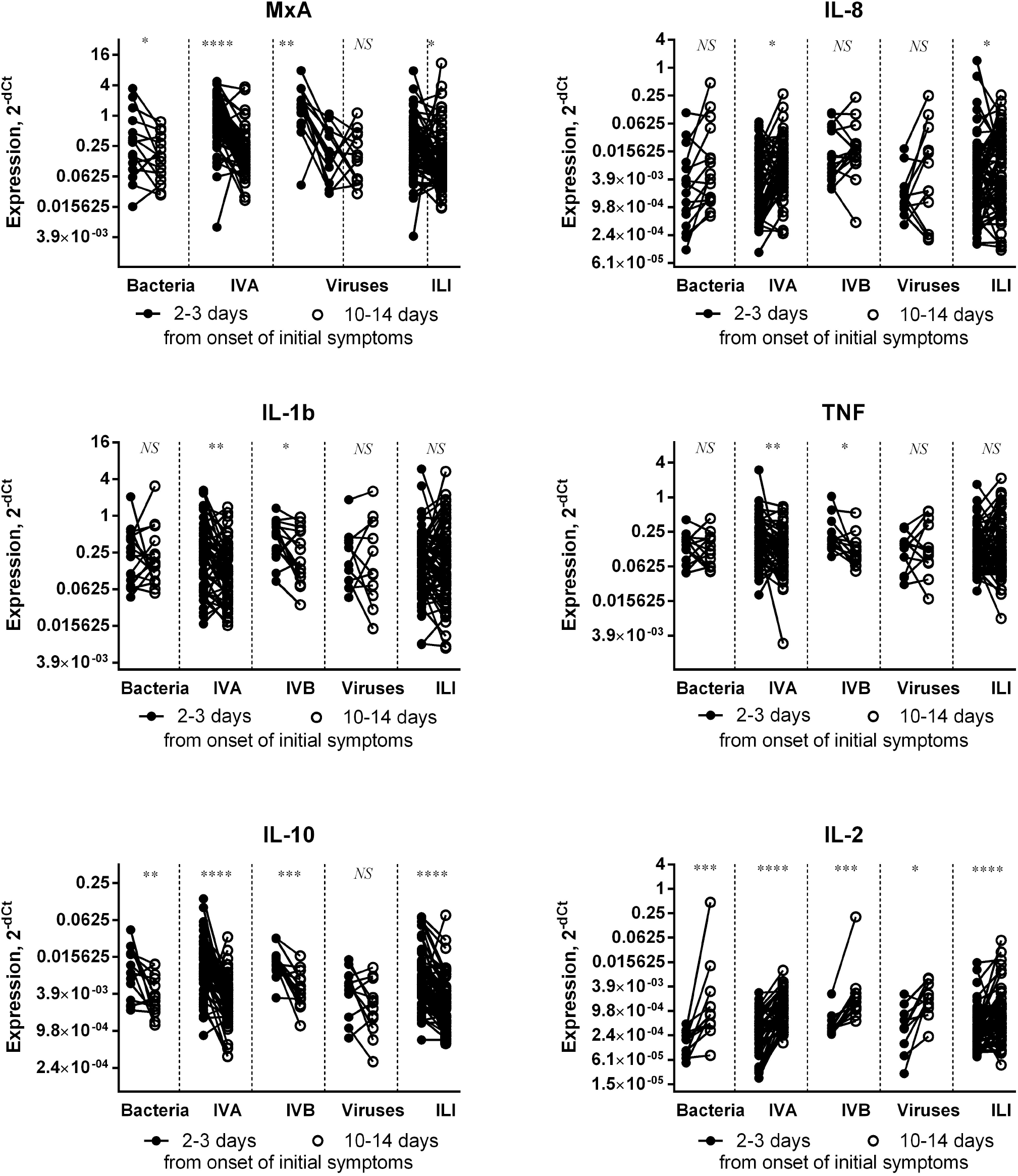

In almost all non-control study groups (with the exception of the viral groups ‘Influenza Viruses’ and ‘ILI Other Viruses’), decreases in the anti-inflammatory cytokine IL-10 were also observed by days 9-10 (Table 1). It is noteworthy that, in all patient groups, there was a significant increase in IL-2 by days 10-14 of illness compared with the acute phase of infection. Earlier, on days 3-4 of illness, patient IL-2 expression levels did not differ from the healthy volunteer group. The increase in IL-2 mRNA is probably due to the development of T cell immune memory. Patient WBC IL-18 levels remained stably elevated, even on days 10-14, in patients diagnosed with influenza, with levels similar to those measured on days 2-3 of illness. Interleukin-4 levels on days 10-14 of illness did not change relative to those on days 2-3. Moreover, when comparing IL-4 over time with the healthy volunteer group, no significant differences were noted. Thus, it can be assumed that our study did not reveal any significant differences in WBC IL-4 expression in pathological conditions caused by ILIs.

## Discussion

This work was devoted to analysis of cytokine expression in patients with acute, seasonal, respiratory infections from etiological agents circulating in 2018-2019. The study was initiated before the appearance and spread of COVID-19 disease, caused by the SARS-CoV-2 virus, in Russia. To assess the potential of individual pathogens to provoke systemic inflammatory response syndrome, we studied mRNA levels at the systemic level: analysis of patient peripheral circulating WBCs. Most of the infections we identified in patients were of a viral nature and were directly caused by influenza viruses (A or B). As a control group, 30 volunteers (men and women) without chronic illness were selected; they were free of infectious illness at the time of sampling (and in the month prior).

According to our data, infection of the respiratory tract with viral or bacterial pathogens induced the expression of cytokines IL-1b, TNF-α, IL-8, IL-18, IFN-λ, and MxA in patient WBCs on days 2-3 of illness. The production kinetics of these pro-inflammatory markers at the systemic level (WBC expression) were generally similar in patients with various pathogens. However, the greatest (and most significant) changes in expression were caused by influenza A and B viruses. Expression of the proinflammatory cytokines IL-1b, TNF, and MxA had significantly decreased by the time of recovery (days 10-14). Regarding IL-6 expression, we found that significantly increased WBC mRNA levels were seen only with infections caused by viral pathogens or in the ‘infection of undetermined etiology’ group. Bacterial infections did not cause significant changes in IL-6 expression relative to the healthy volunteer group. Interestingly, our results are consistent with those obtained by other researchers: respiratory viral infections in obstructive pulmonary disease induce increased serum IL-6 production compared to bacterial pathogens [Zheng et al., 2017].

In a study of IL-6 expression in viral and bacterial diarrhea [Xu et al., 2018], it was also shown that PBMC IL-6 expression (mRNA and protein) were higher in the viral infection group than in the bacterial infection group. IL-6 plays an important role in cytokine release syndrome. The degree of IL-6 expression change can likely be a criterion for determining the potential of a pathogen to provoke hypercytokinemia development. On the other hand, the absence of significant changes in IL-6 expression in the ‘bacterial’ patient group may be due to ongoing antibiotic therapy [Holub et al., 2013].

Observed IL-8 expression values were higher than normal in all patient groups during acute phase illness, regardless of the pathogen. This indicates the presence of a pronounced inflammatory response. Only with type A influenza, however, was there a tendency towards increasing IL-8 in the early recovery period, relative to the acute phase. The significance of such changes was low (p-value <0.05); presumably, a significant contribution to these changes was made by cases of complicated influenza.

In addition, all patients showed significant increases in IL-10 expression. Systemic IL-10 production is a marker of anti-inflammatory response [de Waal Malefyt et al., 1991]. IL-10 limits inflammatory reaction duration and provides a system of negative regulation of the inflammatory response. This negative feed-back loop limits the development of hypercytokinemia in patients. It is also interesting that, by 10-14 days after the onset of illness, almost all patient groups showed decreased IL-10 expression, which probably reduces the risk of immunoparalysis or the development of opportunistic infection [Saraiva & O’garra, 2010].

It is known that with an adequate acute immune response, induction of IL-10 does not affect viral clearance. However, sustained expression during primary or secondary immune responses may contribute to virus persistence or the development of chronic infection [Rojas et al., 2013]. The delicate balance, between limiting the inflammatory response and IL-10-mediated immune regulation required for T-cell homeostasis and host tissue defense, can be compromised by viruses. Despite the fact that in the viral infection patient group there was also a tendency towards a decrease in IL-10 expression by the time of recovery, statistical analysis showed that such changes were not significant. Perhaps this is due to: a strong heterogeneity of pathogens in this group; a relatively small sample of patients for specific viruses; or presence of latent viral infections in patients, producing viral IL-10 homologs.

Curiously, we did not observe any changes in systemic expression of another anti-inflammatory cytokine, IL-4, in patients (neither on days 2-3 of illness, nor on days 10-14). Despite the fact that changes in patient WBC IL-2 levels were not significant on day 2-3 of illness, all patient groups showed a significant increase in expression by the time of recovery. Numerous studies have shown that IL-2 is responsible for the clonal expansion of antigen-selected CD4^+^ and CD8^+^ T cells [Nelson, 2004; Robb et al., 1981]. It also enhances B-cell growth and synthesis of immunoglobulins [Butler, 1998; Arima et al., 1986]. With a high degree of probability, it can be assumed that induction of IL-2 production by WBCs on days 10-14 after illness is due to the development of an adaptive immune response and the formation of immunological memory in patients.

## Conclusion

The vast majority of epidemics in the 21st century, including the current COVID-19 pandemic, have been driven by respiratory viruses. Obviously, a key factor for development of severe pathology can be not so much the pathogen itself but the nature of the host organism’s immune response to it. Current research points to cellular and molecular contributions to cytokine storm phenomena in various disease states. Analysis of the nuances of systemic cytokine production provides several benefits. This and further data, specific for certain viral and bacterial pathogens, will likely make it possible to: assess the risks of developing hypercytokinemia during respiratory infection with agents circulating in the human population; and to rationally predict the pathogenicity and virulence of circulating threats.

## Data Availability

Informed consent was obtained from all donors and patients who provided research materials.

## Author Contributions

The authors contributed to the work, as follows: A.A.L. and A.A.T. performed registration of biological samples and isolation of WBCs; K.I.L. and M.A.E. performed RNA extraction experiments; I.L.B., M.A.E., M.A.P., and S.A.K. performed RT-PCR analysis; M.A.P. performed analysis of RT-PCR data and wrote all manuscript sections; S.A.K. and A.V.V. provided funding acquisition, supervision of research; M.A.P. wrote the first draft of the manuscript; E.S.R. prepared the final draft of this paper (translation, editing). All of the authors have read and approved the final manuscript.

## Ethics

The study and its protocols were approved by the Smorodintsev Research Institute of Influenza institutional Ethics Committee (protocol № 108, dated September 03, 2018). Serum specimens used for isolation of white blood cells (control group) were provided by the Blood Transfusion Center, Research Institute of Hematology and Transfusion Science (contract number 128_15092017). Informed consent was obtained from all donors and patients who provided research materials. All biological experiments were performed in accordance with the relevant guidelines and regulations.

## Abbreviations

IVA: influenza A virus(es)
IVB: influenza B virus(es)
RSV: human orthopneumovirus(es)
HMPV: human Metapneumovirus(es)
HPIV: human parainfluenza virus(es)
HCoV: human Coronavirus(es)
HRV: human Rhinovirus(es)
HAdV: human Adenovirus(es)

